# Impact of Dutasteride on PSA Kinetics and Time to Progression in Men with PSA-only Recurrent Prostate Cancer

**DOI:** 10.1101/2021.05.03.21256519

**Authors:** Charles E Myers, Michelle S McCarthy

**Author notes:** Corresponding Author. Foundation for Cancer Research and Education, P.O Box 746, Earlysville, Virginia 22936, USA.

## Abstract

**Background:** AVODART After Radical Therapy For Prostate Cancer Study (ARTS) reported dutasteride reduced PSA doubling (PSADT) at two years by 66% and disease progression by 59%. The durability of the cancer control is unknown.

**Objective:** Explore the impact of dutasteride on PSADT and time to progression in men with PSA-only recurrent disease.

**Design, Setting and Participants:** Retrospective examination of the impact of dutasteride on PSA kinetics and time to progression with PSA-only recurrent prostate cancer.

**Intervention:** Dutasteride daily.

**Outcome, Measurement and Statistical Analysis:** Change in PSA over time was determined by linear regression of natural log of PSA versus time. The slope of that curve was used as a measure of exponential PSA progression. Impact of dutasteride on slope was analyzed using Wilcoxon signed-rank two-sided test. Time to progression was analyzed using Kaplan-Meier, univariant and multivariant Cox regression.

**Results and Limitation:** Compared to men with BPH, patients showed little change in PSA during the first 3 months. Thereafter, the PSA resumed an exponential increase. PSADT was 10.3 months pre dutasteride and 24.8 months post dutasteride. Multivariant analysis showed a strong correlation between post dutasteride PSA kinetics and time to progression, with close to 50% relapse free at 10 years. Post-dutasteride, patients with PSADT >9 months had significantly better survival.

**Conclusions:** Dutasteride slows PSADT in PSA-only recurrent prostate cancer. This decline correlated with time to disease progression with close to 50% progression free at 10 years.

**Patient Summary:** Men with PSA-only recurrent prostate cancer on dutasteride experience a significant drop in the rate at which the PSA increases. If the PSA does not double between months 3-12, they are likely to remain free of metastases for many years.

## Introduction

Optimal treatment for PSA-only recurrent prostate cancer remains controversial^1,2^. This is especially so for patients with a PSADT > 9 months, who can remain free of detectable metastases for greater than 10 years^3^. While these patients initially respond to standard hormonal therapy, a survival benefit to early treatment has yet to be proven and the morbidity associated with long-term treatment is considerable^4^.

This has led to a search for treatments that further slow the growth of these indolent cancers^5,6^. Agents that have been tested suppress cancer progression include those that trigger an immune response to the cancer or act biochemically to hinder cancer growth^5,6^. As these patients do not have detectable cancer, therapeutic effect is measured by a change in exponential increase in PSA over time, most commonly expressed as the PSADT^7^.

Dutasteride is one drug that has been reported to improve cancer control in this setting. The Avodart After Radical Therapy for Prostate Cancer Study (ARTS) trial was a randomized controlled study that found dutasteride reduced the risk of PSA doubling at two years by 66% and the risk of disease progression by 59%^8^.

However, there are concerns about the use of PSA kinetics in patients on dutasteride. In patients with BPH, dutasteride commonly causes an initial rapid fall in the PSA level^9,10^. If a similar rapid decline in PSA occurred after dutasteride administration in men recurrent after initial surgery or radiation, then it would complicate calculation of PSA doubling time.

We demonstrate that on dutasteride, PSADT can be accurately determined starting 90 days after the start of therapy. The PSADT went from 10.3 months pre-dutasteride to 24.8 months post-dutasteride. Furthermore, the reduced PSADT correlates well with time to progression.

## Methods

The primary goal of this study was to examine the impact of dutasteride on PSA kinetics in men with PSA-only recurrent prostate cancer. We then planned to examine whether the impact of dutasteride on PSA kinetics correlated with time to progression. We searched our clinic database to identify patients with sufficient pre- and post-dutasteride PSA information to assess the impact of the drug on the PSA kinetics. To that end, we applied the following criteria^7^:

1. PSA-only recurrent prostate cancer after initial treatment with curative intent who were placed on dutasteride as their only cancer treatment;
2. Must not have received dutasteride previously;
3. Minimum 3 PSA values in the year prior to the initiation of dutasteride;
4. Minimum of 4 PSA values in the first year on dutasteride;
5. PSA detectable and increasing prior to initiation of dutasteride and still detectable for 180 days post dutasteride. The latter requirement was needed to have sufficient information to calculate the change in PSA kinetics, but did exclude a number of patients whose PSA rapidly became undetectable;
6. PSA values from a single testing laboratory and from the actual lab report rather than patient word of mouth.

### Dutasteride Dosing

Dutasteride administration is commonly 0.5 mg once a day, but clinical trials have demonstrated safety at higher doses^11^. We started patients at this standard dose and schedule. As dihydrotestosterone (DHT) was monitored, the dosing was increased or decreased to keep the DHT at or below 5 ng/dL. Twenty-eight patients remained at one 0.5 mg capsule a day. Five patients were on a reduced dose within the first year: one to 1 capsule every other day, three to 1 capsule 3 times a week and one to 1 capsule twice a week Three patients had to increase their dose within the first year: one to 2 per day and two to 4 per day.

Progression was defined as either the start of any additional treatment for prostate cancer or the end of follow up associated with referral to another physician or the closure of the clinic.

### Statistical Considerations

All statistical analysis was performed using XLSTAT by Addinsoft statistical and data analysis solution (2019.4.2 Build 63762), New York, USA.

PSA kinetics were analyzed using linear regression analysis of natural log of PSA (ln PSA) versus time^7^. The PSA slope is represented by the slope of that line. It is common practice to convert the PSA slope to a PSADT, as that metric is convenient for clinical management. However, 15 out of 47 patients showed a negative slope throughout this time period. As calculating a PSA doubling time for a declining PSA has no meaning and is not done, analysis was done using the PSA slope. Where appropriate, PSADT was calculated by dividing the natural log of 2 by the slope.

Because both the pre- and post-dutasteride PSA velocities were not normally distributed, the statistical analysis of these results was performed using the Wilcoxon signed-rank two-sided test.

The association between changes in PSA slope and time to progression was analyzed using the Cox proportional hazard model. First, a range of potential variables were subject to a univariant analysis. In order to obtain the multivariate model, a forward model selection was applied to the set of variables which were significant in the univariate analysis. Additional covariates were included in the model if they significantly improved the model’s likelihood ratio (p < 0.1). Kaplan-Meier estimator was used to analyze time to progression.

## Results

### Patient Characteristics

The first patient started on dutasteride 5/7/03. Follow up ended on 9/20/17 with the closure of the clinic.

Our analysis involved 47 unique patients (Table 1). The median age was 58, with a range of 43 to 74.

**Table 1.**
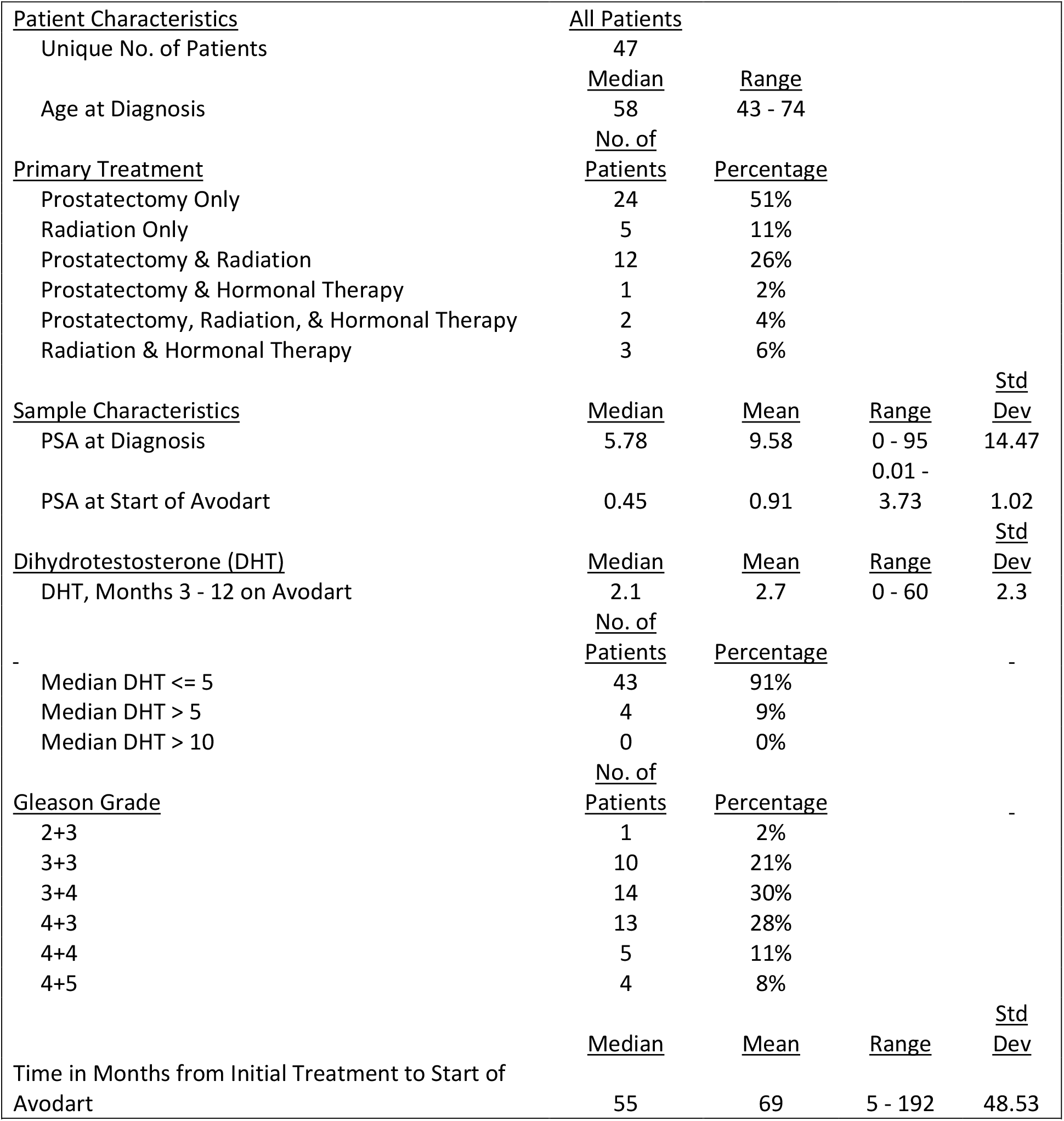
Patient Characteristics

Prostatectomy alone was the most common treatment at 51% (twenty-four individuals). The second largest group had prostatectomy followed by radiation at 26% (twelve individuals). Five patients (11%) had radiation alone. One patient had prostatectomy + hormonal therapy, two had prostatectomy + radiation therapy + hormonal therapy and three had radiation therapy and hormonal therapy. Thus, a total of six (12.7%) had hormonal therapy in combination with either radiation or prostatectomy.

Gleason grades were predominantly lower than eight with thirteen at 3+3, fourteen at 3+4 and thirteen at 4+3. This accounted for 78.4% of the patients. A total of nine had higher Gleason grades with four at 4+4 and five at 4+5

The median PSA at the start of dutasteride was 0.4 ng/ml, with a range of 0.10 to 3.80 ng/ml. Patients with a PSA below 0.2 ng/ml were required to have an exponentially increasing PSA documented by ultrasensitive PSA assay.

The mean and median DHT were 65.3ng/dL and 66.0 ng/dL, respectively, with a standard deviation of 7.2 ng/dL. These values are well within the published normal ranges for males in this age group. Six patients had previous hormonal therapy and three had reduced DHT levels of 31, 17 and 9 ng/dL.

### Dutasteride Impact on DHT

As dutasteride acts by suppressing the conversion of testosterone to DHT, we measured DHT monthly during the first year on this drug. This provided an assessment of patient compliance.

Previous studies have shown that dutasteride takes 3 months to reach steady state concentration and the full impact of dutasteride takes similar time to develop^11^. For this reason, we used a mean and median DHT for months 3-12 as a measure of dutasteride suppression of DHT. The mean and median DHT during this time interval was 2.7 ng/dL and 2.1 ng/dL. Thus, dutasteride administration resulted in a 95.9% or 96.3% suppression using the means or medians, respectively.

We had set a mean DHT of less than 5 ng/dL as the goal of treatment. A total of 43 patients (91%) reached that goal. Four patients (9%) had mean DHT between 5 ng/dL and 10 ng/dL, while no patient had mean DHT above 10 ng/dL ng/dL. Thus, all patients had at least an 84.7% suppression of DHT during this time period and indicate adequate patient compliance with the treatment program.

### Impact of dutasteride on PSA kinetics

As previously mentioned, it takes 3 months for dutasteride to reach steady state levels and this is apparent in the PSA kinetics. The Waterfall plot of PSA change over the first 90 days shows limited impact of the drug on levels of this marker (Figure 1). For this reason, we used the PSA from 3 months to 12 months after the initiation of dutasteride to determine the impact of the drug on PSA kinetics.

**Figure 1.**
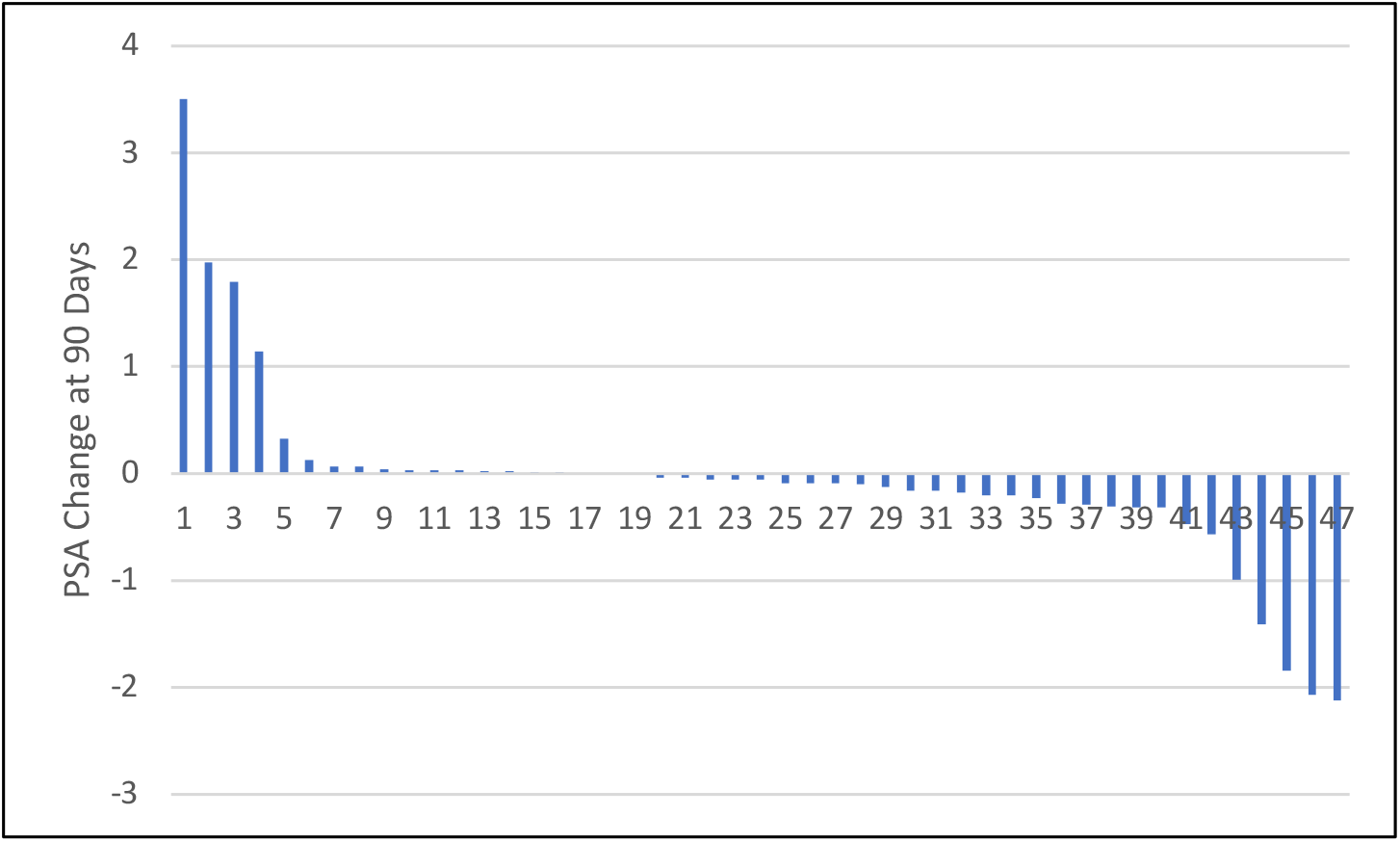
Waterfall plot of the change in PSA over the first 90 days following the initiation of dutasteride.

We calculated the PSA slope for both pre and post dutasteride intervals. The mean slope pre-dutasteride was 0.067 +/-0.073 SD ln PSA/month compared with 0.028 +/-0.062 SD ln PSA/month post dutasteride, for a 58% decrease in PSA slope.

These numbers represent a PSADT of 10.3 months pre dutasteride and 24.8 months post dutasteride. Figure 2 shows a Waterfall plot of pre-post slope.

**Figure 2.**
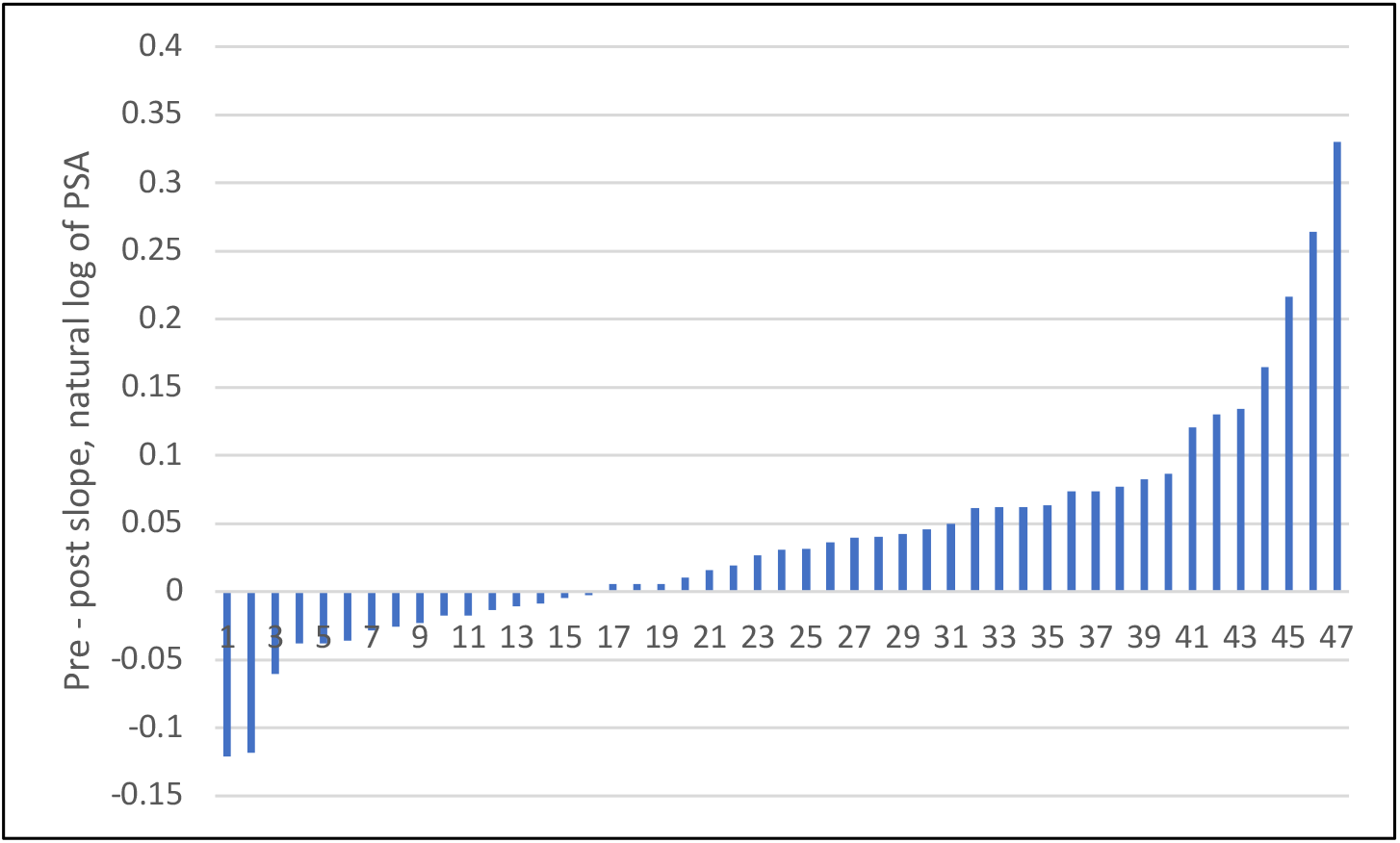
Waterfall plot pre minus post slope, nl PSA, ng/ml versus time, months.

Both the pre and post dutasteride slopes results failed the test for normality (Shapiro-Wilk). Therefore, the statistical significance of the results was analyzed using the Wilcoxon signed rank two tail test and yielded a p= 0.001.

### Post-Dutasteride PSA Kinetics and Time to Progression

We next examined the relationship between the change in PSA kinetics and time to progression on dutasteride. Figure 3 left panel shows the Kaplan-Meier plot for all patients. The median time to progression was 135 +/-8.5 months SD.

**Figure 3.**
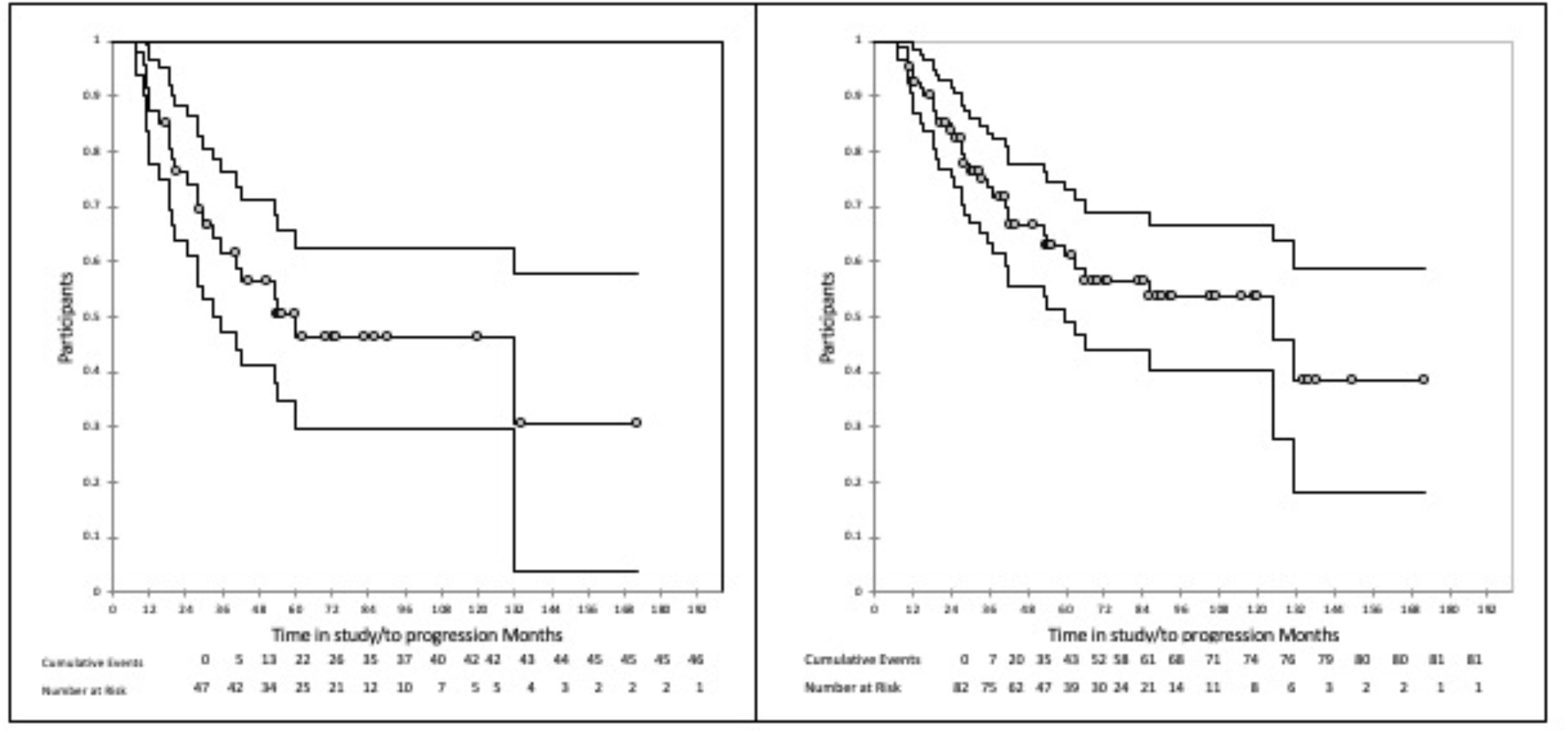
Kaplan Meier plot of time to progression on dutasteride. The panel on the left shows the result for the patients in this study. The panel on the right shows the result for the larger group of patients from which the study group was selected.

Patients in this paper were selected from a larger group of 82 patients with recurrent prostate cancer treated with dutasteride, using the selection criteria listed in the Methods section. Figure 3 right panel shows a Kaplan Meier plot of those patients with a mean survival time of 93.6 +/-7.3 months SD.

Next, we did univariant Cox proportional hazard model analysis to evaluate the contribution of other variables to the outcome. The variables tested include pre-dutasteride PSA slope, total PSA change over the first 12 months on treatment, post-dutasteride PSA slope, pre-post dutasteride PSA slope, PSA change in first 90 days, prostatectomy only, radiation only, PSA at time 0, time from initial treatment, Gleason 3+3 or 3+4 yes or no, and PSA slope negative (yes or no). On univariant analysis, only the four variables were statistically significant: PSA change over the first 12 months (p=0.005), PSA slope post-dutasteride (p=0.003), the difference between the pre- and post-dutasteride PSA slope (p=0.008), and PSA change in the first 90 days (p=0.018). Prostatectomy yes or no, was just outside of significance with p=0.064. All of the other variables had p>0.2.

The four variables significant on univariate analysis were then investigated in a multivariate Cox Proportional Hazards model with forward model selection. Two variables emerged from this analysis: post dutasteride slope and PSA change at 90 days. The resulting model fit the data with a p<0.0001. Below are the regression coefficients for the two variables:

| Variable | Value | Standard error | Wald Chi-Square | Pr > Chi<sup>2</sup> | Hazard ratio | Hazard ratio Lower bound (95%) | Hazard ratio Upper bound (95%) |
| --- | --- | --- | --- | --- | --- | --- | --- |
| Post Dutasteride Slope | 10.310 | 4.407 | 5.473 | 0.019 | $30 \times 10^3$ | 5.326 | $169.2 \times 10^9$ |
| PSA Change at 90 Days | 0.492 | 0.312 | 2.488 | 0.115 | 1.6 | 0.888 | 3.011 |

PSADT is an established prognostic marker in men with cancer recurrent after initial surgery or radiation. Generally, PSADT slower than 9-12 months have been used to identify men with a more favorable prognosis^3,12^. With this in mind, we looked at the association between PSA doubling times of slower than 9 or 12 months during months 3-12 and time to progression. This analysis is shown in Figure 4. Both 9 and 12 months would serve to identify a group of patients with greater than a 50% of still being on dutasteride at 10 years. Also, in both cases the p value was significant at 0.002. This suggests that the PSADT during the first year might be useful in identifying those most likely to have durable disease control on dutasteride.

**Figure 4.**
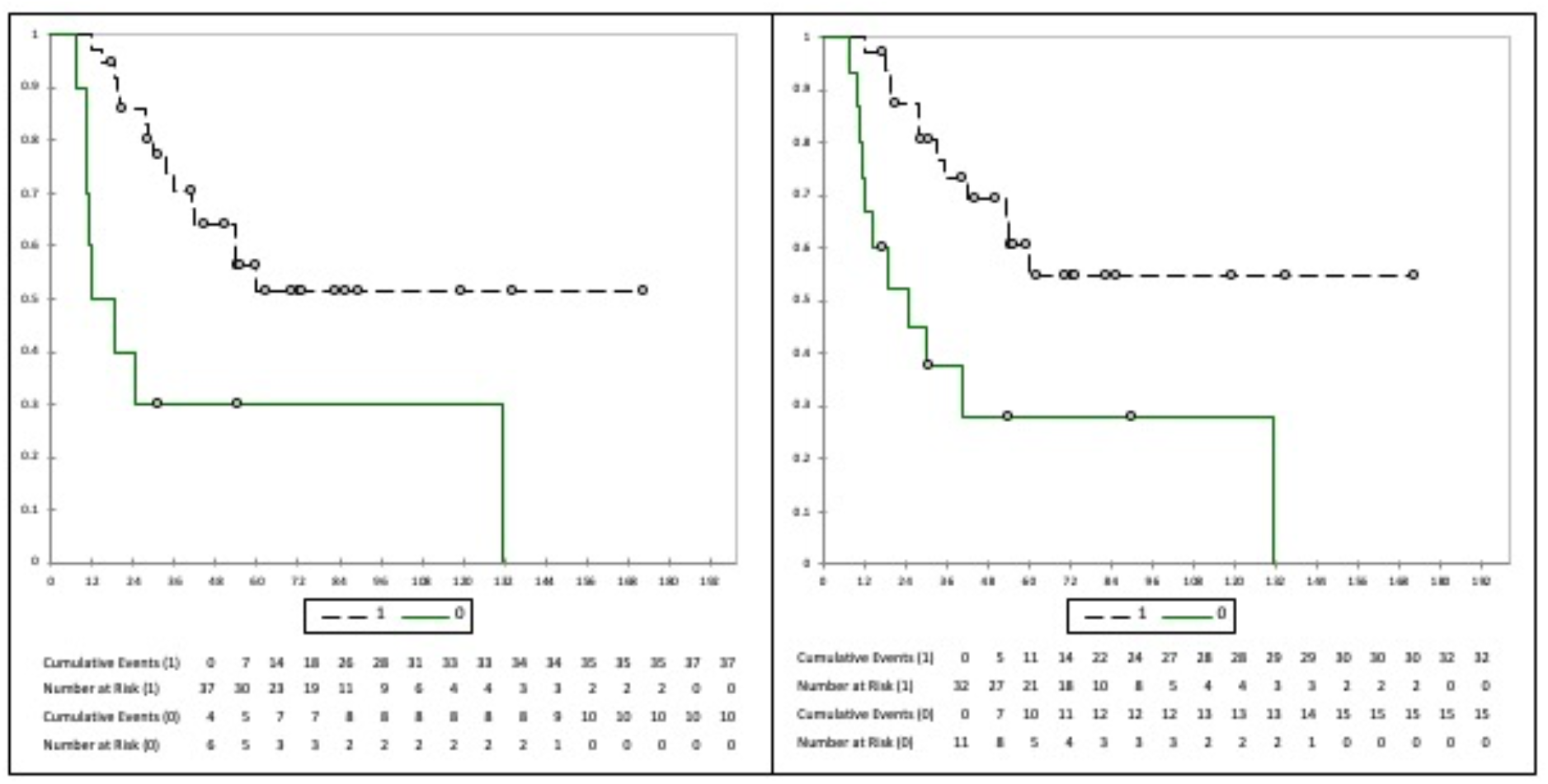
Kaplan Meier plot of time to progression for those whose did (1) or did not (0) double in 9 months (left panel) or 12 months (right panel). For both panels, p was 0.002.

## Discussion

There have been concerns about the validity of PSADT in patients on dutasteride. However, in contrast to men with BPH, we found little net change in PSA during the first 90 days on dutasteride. As far as we can determine, this is the first report of the marked difference PSA in kinetics on dutasteride between men with BPH versus those with recurrent prostate cancer.

We found that dutasteride resulted in a 58% reduction in the PSA slope that was statistically significant(p=0.001). These results are consistent with those from the ARTS trial as they reported 59% of patients free of progression at 2 years and our Kaplan-Meier plots show a plateau close to 50% out to 10 years^8^. These results suggest that dutasteride may offer durable cancer control to a portion of patients with recurrent disease. This would need confirmation in a randomized controlled trial.

We looked at early events that might help identify patients likely to benefit from long-term dutasteride treatment. On multivariant analysis, no pretreatment characteristics proved to be statistically significant. Instead, post dutasteride PSA kinetics appeared to be the major determinant of time to progression.

PSADT slower than 9-12 months is widely used to identify patients with a favorable prognosis. It appears that this guideline may also be applicable to patients with PSA-only recurrent disease on dutasteride as PSADT slower than 9 months was associated with an excellent prognosis.

There is a growing list of agents that have been shown to slow PSA kinetics in this group of patients^5,6,13^. Among such agents, dutasteride has many advantages. It can be given once a day. Because it has such a long half-life, missing a dose will have little impact on DHT suppression. There is extensive safety information and the risk of severe toxicity is quite low, even after many years of use. It has no major interactions with other drugs commonly used in men over 60 years of age, such as drugs for cardiovascular disease or diabetes. Dutasteride costs $0.27-0.33 a day and it is widely available.

One key reservation is that the Clinical Research Study To Reduce The Incidence Of Prostate Cancer In Men Who Are At Increased Risk (REDUCE) noted an increase in the number of high-grade cancers in the drug arm compared to the placebo^14^. The Prostate Cancer Prevention trial that looked at finasteride showed a similar increase in high-grade prostate cancer^15^. The meaning of these findings remains controversial, but it has not been shown that the appearance of these high-grade lesions had any negative impact on clinical outcome. Furthermore, the REDEEM trial evaluating the role of dutasteride in active surveillance found no evidence of an adverse impact of dutasteride^16^. The same was true of the ARTS trial^8^.

## Conclusions

Dutasteride warrants more a detailed evaluation as an agent to slow progression in patients with PSA-only recurrent disease.

## Data Availability

Study data is available upon request

## Acknowledgement

Jean Paul Maalouf, Data Science Consultant, Addinsoft provided advice on the performance of the Cox model analysis. **Paul Schellhammer**, MD. Virginia Prostate Center, Eastern Virginia Medical School in Norfolk, Virginia for suggesting several significant revisions in the initial draft.

## Notes

FUNDING This study was funded by Foundation for Cancer Research and Education, Holewinski Family Foundation Inc and Schellhammer Urological Research Foundation.

### Competing Interest Statement

The authors have declared no competing interest.

### Funding Statement

This study was funded by Foundation for Cancer Research and Education, Holewinski Family Foundation Inc and Schellhammer Urological Research Foundation.

### Author Declarations

Integreview IRB, 3815 S. Capital of Texas Highway, Suite 320, Austin TX 78704, 512-326-3007 | Fax: 512-593-6040 www.integreview.com, reviewed our protocol and designated that study was exempt under category 4.

